# COVID-19 Outbreak Situations in Bangladesh: An Empirical Analysis

**DOI:** 10.1101/2020.04.16.20068312

**Authors:** Md Hasinur Rahaman Khan, Ahmed Hossain

## Abstract

COVID-19 disease, as popularly known as Coronavirus 2019 disease, has been emerged from Wuhan, China in December 2019 and now is a pandemic for almost every nation in the earth. It affects every country without considering country’s race, nationality and economic status. This paper aims at analysing primarily the current situations of Bangladesh and predicting infections and deaths for moderated term intervals by a proposed projection technique called Infection Trajectory-Pathway Strategy (ITPS) and for short term intervals prediction for total infections, deaths along with total number of severe patients and Intensive Care Unit (ICU) patients by polynomial regression modeling approach. Since April 7, Bangladesh has started to face critical situations as the number of infections has accelerated very fast in the following days. However, the fatality rate decreases considerably from 15.7 on April 1 to 4.9 on April 14, which is still high among the south asian countries. Of the 1012 cases reported on April 14, almost 70% are the male, 42% are from the capital Dhaka. We have found that the potential pathway of infections for Bangldesh would be the similar pathways that are experienced by Austria, Netherlands, Israel, France and United Kingdom. These countries are ahead a number of weeks and days in terms of infection cases since their 100-th confirmed cases. Our proposed projection method ITPS suggests that by May 10, Bangladesh will cross 12000 incidences and 720 deaths which, by May 16 will be 27000 and 1644 respectively. On the other hand, the regression model suggests that by the end of April, total number of infections, deaths, severe patients and ICU patients will be 5780, 347, 775, and 694 respectively. This study will be favorable for the administrative units of Bangladesh to plan for the next few weeks and to consider various aspects related to the control of COVID-19 outspread in Bangladesh.

## 1 Introduction

Severe acute respiratory syndrome coronavirus (SARS-CoV-2) caauses coronavirus disease 2019 which is an infectious disease and was first identified in December 2019 in Wuhan, the capital of China’s Hubei province. This disease spreads first in Wuhun to great extent in December 2019 and January 2020, then spread globally since February resulting in the ongoing 2019-20 coronavirus pandemic. For most people, COVID-19 infection will cause mild illness like fever, cough and shortness of breath. However, it can make some people very ill and it can be, in its worst scenario case, fatal. Older people, and those with pre-existing medical conditions (such as cardiovascular disease, chronic respiratory disease or diabetes) are at risk for severe disease (WHO, 2020). Other common symptoms maay include fatigue, muscle pain, diarrhea, sore throat, loss of smell and abdominal pain (Wikipedia, 2020).

Bangladesh found first coronavirus cases on 8 March. The first three coronavirus cases were confirmed by the IEDCR with at a press conference (IECDR, 2020). The cases included two men and one woman, who were aged between 20 and 35. Of them, two men were Italy returnees and the woman was a family member of one of these two. Approximately about 111 tests were conducted on that day in Bangladesh. On March 16, the country detected three more cases of COVID-19, taking the total number of infected people to 8. Bangladesh have first death due to coronavirus on March 18. A 70-year-old man died of the disease. Pandemic forced Bangladesh like other countries in the world to adopt several measures like compulsury lockdowns, home quarentine, social distancing and local or international flight bans etc. for slowing down the spreading. Bangladesh followed shutting down schools and colleges on March 18 and one week later from March 26 the all offices remain close resulting national lockdown. As of April 14, number of COVID-19 infections is 1012 and that has already spread in 38 districts of 64 districts (IECDR, 2020).

But for people in one of the densest countries on earth, it is difficult task to maintain social distancing, despite closing of educational institutes, offices, and markets may contribute considerably to reducing spread, while commuting in crowded public transport or even living in cheek by jowl urban slums. Besides, public healthcare system in Bangladesh is not sufficient, rather is overburdened. According to World Bank data (WB, 2020), Bangaldesh in 2015 has 0.8 hospital beds for every 1,000 people; by way of comparison, the India has 0.7 (2011), the Pakistan has 0.6 (2012), the SriLanka has 3.6 (2012), US has 2.9 (2012) while China has 4.2 (2012) beds per 1,000 people. Currently, hospitals in Bangladesh have total 1,169 ICU beds of which 432 are in government hospital and the remaining 737 in the private hospitals against a population of 170 milion (“The Daily DhakaTribune”, March 21, 2020). Total number of ICU beds in a hospital should be between 5% and 12% depending on the care given by the hospital [(Kennedy & Pronovost, 2006), (Nafseen, 2018)]. In 2017-18, the total number of beds in hospitals were 1,27,360, among them 48,934 were in governmental hospitals and 78,426 were in private hospitals [(Nafseen, 2018), (Ministry of Health and Family Welfare, 2017)]. Currently, Bangladesh has total 141,903 hospital beds (“The Daily DhakaTribune”, March 21, 2020), which is 0.84 beds for every 1,000 people.

Amid of the scarcity in basic health care facilities, has Bangladesh acted accordingly to cope up with the ongoing COVID-19 threats despite facing the underlying economic threats and uncertainty. If so, then how much preparation the country need to take for facing the potential management crisis. Is the country alien to viral outbreaks like COVID-19, having suffered counntry’s worst recorded case of Dengue outbreak in 2019 and at the same time despite having a three-month head start since the outbreak began in China? This paper will help to know the answers to these questions in both directly and indirectly.

As of today, we have found very few research works on COVID-19 that have been conducted using Bangladesh data. Recently, Islam et al. (2020) proposed a model to measure the risk of infectious disease and predict the risk of COVID-19 transmission using Bangladesh data along with other four countries-United States, Australia, Canada and China. Paul et al. (2020) proposed a SEIR epidemic model that accommodates the effects of lockdown and individual based precautionary measures and used it to estimate model parameters from the epidemic data for three south asian countries-Bangladesh, India and Pakistan. Paul et al. (2020) used Bangladesh data up to 2nd April, 2020 by which, Bangladesh has only 95 infection cases and 9 deaths. However, their prediction model for Bangladesh may not give reasonable results because of sufficiently small sample. Mamun and Grith (2020) discussed possible suicide prevention strategies while the first COVID-19 suicide case in Bangladesh took place due to fear of COVID-19 and xenophobia. Neither of the three research works dealt with the analysis of current COVID-19 situations in Bangladesh and to make direct projections for incidence, deaths, hospital ICU beds, number of severe patients etc that are the main goals of this paper. These statistics may help government to take proper preparation to tackle the potential unprecidented situations in Bangladesh.

There is a number of models available in literature to model infectious diseases of which a few models has been used primarily for the countries where number of cases is very high like China, Italy, Spain, UK, Germany and USA. Particularly, a number of study works [Kucharski et al. (2020), Chinazzi et al. (2020), Roosa et al. (2020), Grasselli et al. (2020), Boldog et al. (2020), Hui et al. (2020), Xie et al. (2020), Lourenco et al. (2020), IHME COVID-19 health service utilization forecasting team (2020), Phua et al. (2020) and Phua et al. (2020)] has used various mathematical models to determine the spread of the disease, predict the number of incidence, health care faciities in tackling COVID-19 spread. We will use polynomial regression model to predict the infected people, deaths and other healthcare faciities. However, it was also claimed that Bangladesh’s existing healthcare infrastructure is not very strong as per the WHO guide-lines [9] and in case of community spread, the Bangladesh government may find it difficult to manage the spread in light with the predicted statisstics.

## 2 Data Source

The data used for the current study has been from the Esri Living Atla (Dong & Gardner, 2020). This is the data repository maintained by the Johns Hopkins University [Johns Hopkins University (2020b), Johns Hopkins University (2020a)]. In the current study, we have used other countriy’s (including Bangladesh) temporal data of confirmed and death cases till 14-th April, 2020. For projection we have used only April month’s data i.e. from April 1 to 14 as there is sufficient daily cases from the begining of April. We have also used several other secondary data sources such as the IEDCR (IECDR, 2020), World in Data (Max Roser & Ortiz-Ospina, 2020) and World Bank (WB, 2020).

## 3 Methodology

For basic statistical analysis, we have used the basic statistical tools that include the trend line charts, correlation, t-tests etc. For projection of infections and deaths, we have fitted the incidence and deaths of Covid-19 disease in Bangladesh by higher order polynomial regression. The second order polynomial regression is used with confirming orthogonality that helps to get uncorrelated regression coefficient. The two degree polynomial regression modell is given by

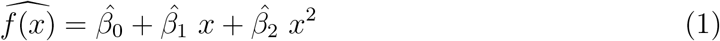

We have obtained also 96% confidence interval estimates. Polynomial regression has been used in forcasting COVID-19 diseases along with for fitting trends by many researches [Pandeya et al. (2020), Johannes (2008), Howard (1943)].

## 4 Analysis

### 4.1 Basic Trends

In Bangladesh, the first COVID-19 case has been detected on March 8, 2020 and the national lockdown was announced after 17 days on 26th March 2020 by the gov-ernment of Bangladesh. This lock down was called well ahead as compared to many other countries including India and Pakistan with a hope of narrow downing the spread of infection. Even before the lockdown majority of the schools, colleges, markets, cinema halls, etc. were already shut down in Dhaka and other parts of the country. Current, the lockdown has been extended until April 26 and government banning all the movements and restricting and urging peole to stay at home. The citizens were allowed to step out only in emergency situations. All these steps were taken in the hope of flattening the curve of infected cases and to limit the exponential growth of the patients in Bangladesh.

The number of infected cases, deaths and active case are reported in Figure 1. There are 1012 infected cases, 46 deaths reported in Bangladesh as on 14-th April 2020 with 91.3% cases being active which is much higher compared to global percentage of active case 70% (Dong & Gardner, 2020). More than 50% cases are from Dhaka division (IECDR, 2020) of which almost 70% are from the capital city Dhaka. The sex ratio (males to females) among the infected population is found 70 to 30 in every 100 cases as of April 12, 2020. Since April 7, the number of infections and deaths has been increased significantly with much higher rates. During April 7 to 14, the average doubling time for infections and deaths is found 3.3 and 4.3 respectively (see Figure 4 (b)).

**Figure 1:**
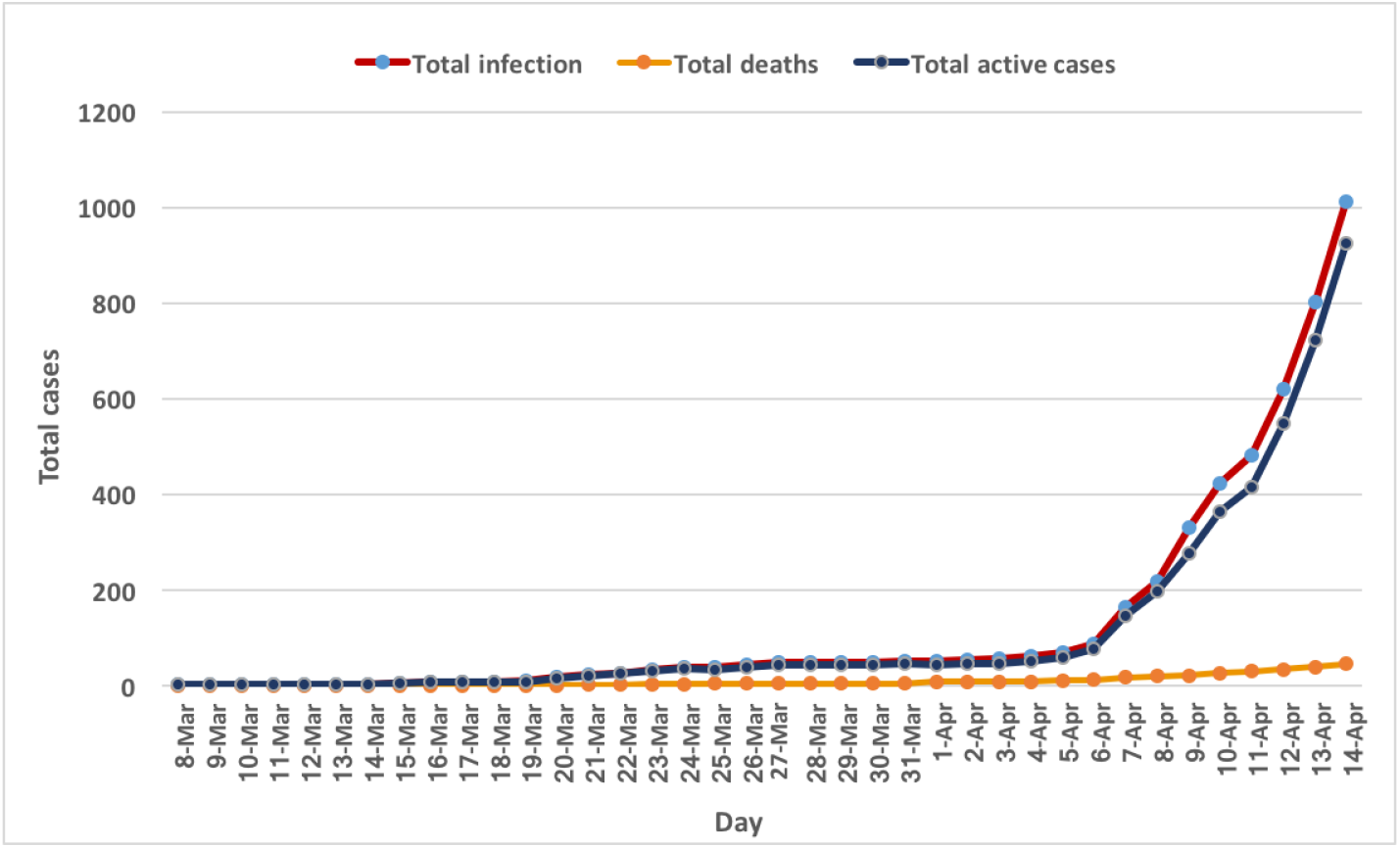
Infected cases, deaths and active cases of COVID-19 in Banglades (as of April 14)

Figure 2 shows the case fatality rate (CFR) and growth rate of infections. CFR is the ratio between the number of confirmed deaths and the number of confirmed cases from COVID-19. Bangladesh had very high CFR rates during April 3 to 5 with average almost 16%. Compared to south asian countries India, SriLanka and Pakistan the CFR rate in Bangladesh is still the highest (see Figure 3). However the current CFR is much lower than the glober CFR which is over 6 as on April 14. In the early days of identification the count was in single digits, so the growth rate was comparatively high, so as the CFR values. But in the weeks that followed, the CFR particularly declined, reaching as low as 4.5% as on April 14, 2020. The growth rate has consistently positive since April 5 and that has been hovering around 25% till April 9.

**Figure 2:**
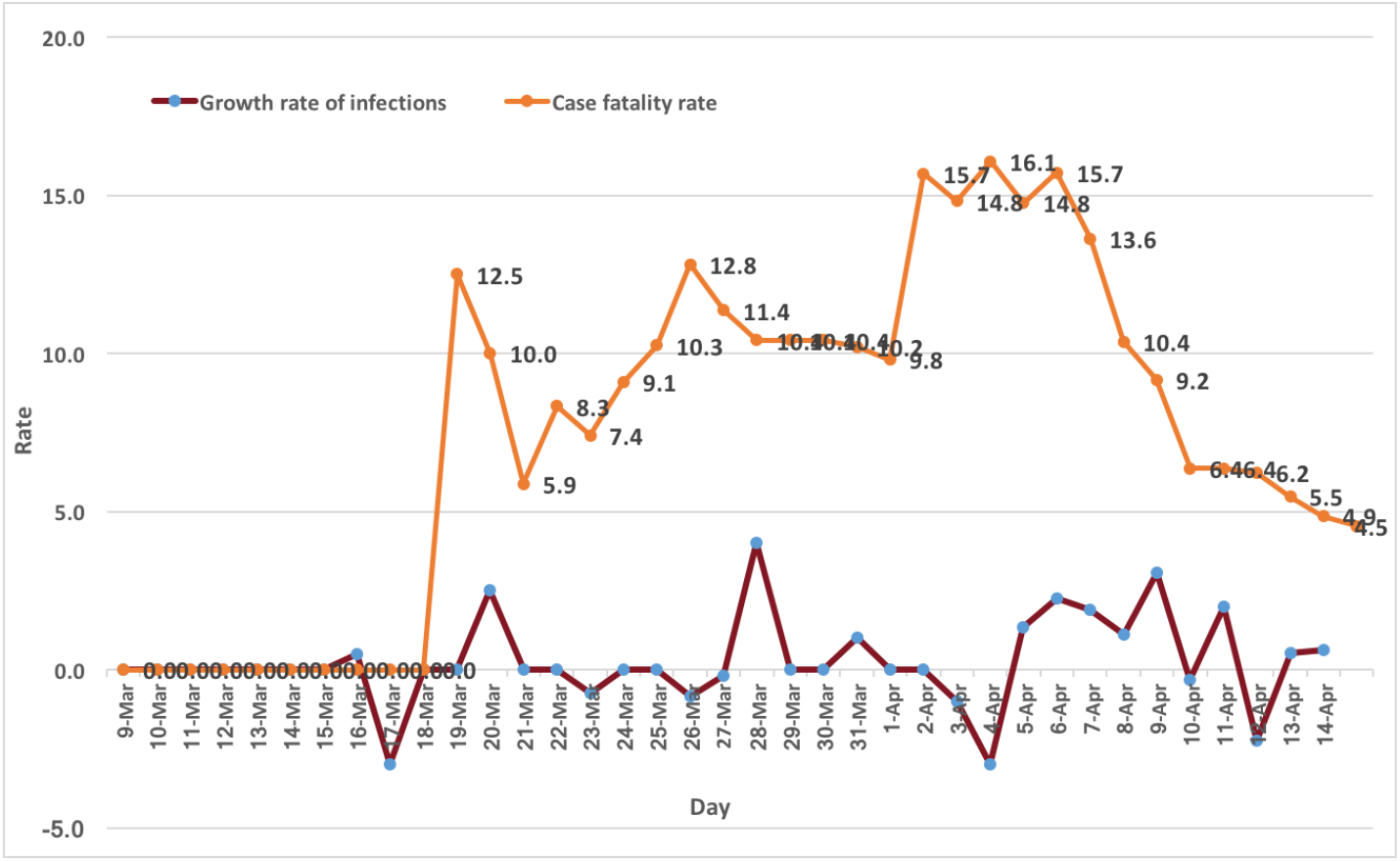
Case fatality rate and growth rate of infections of COVID-19 in Banglades (as of April 14)

**Figure 3:**
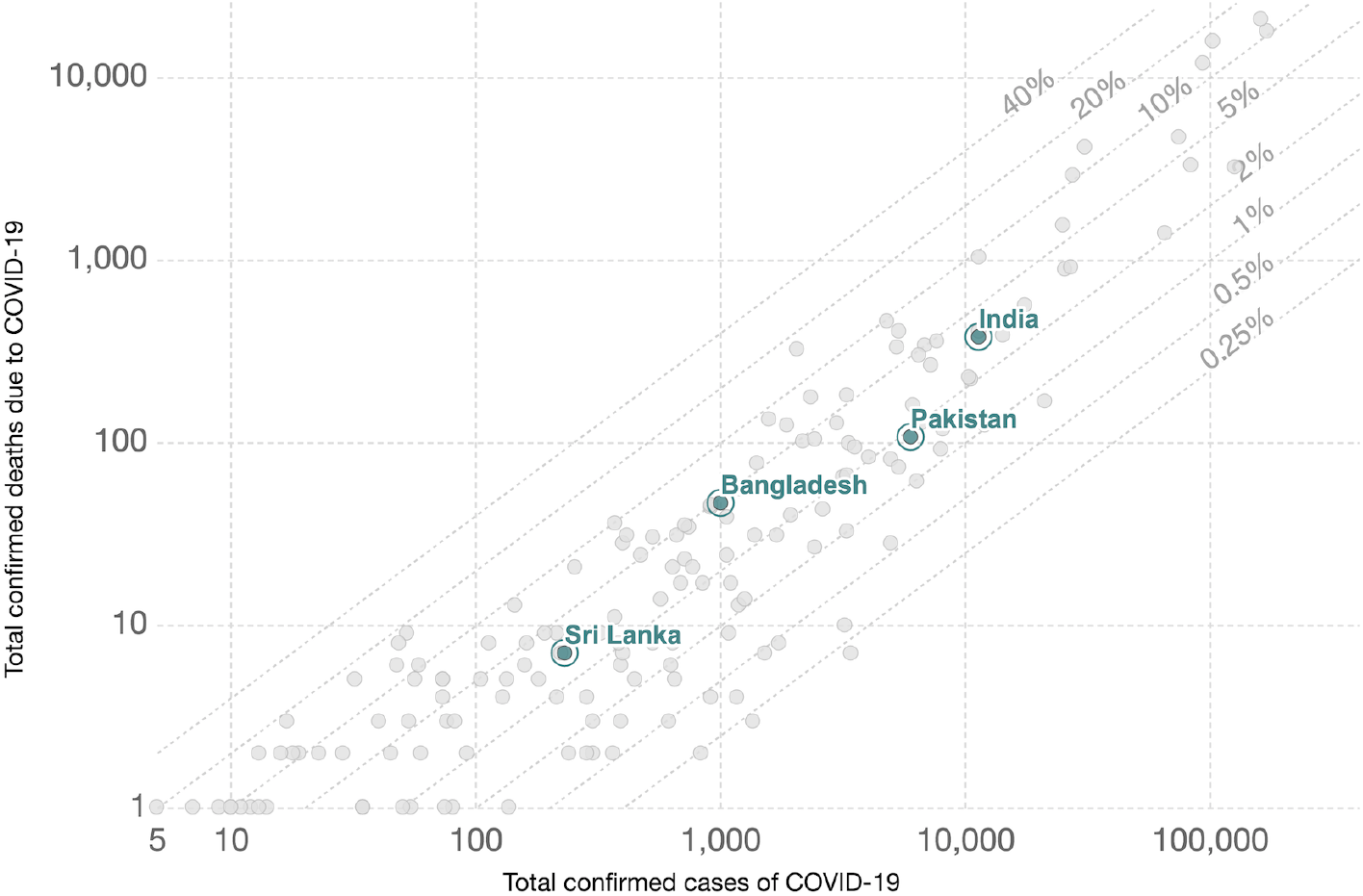
Case fatality rate line- total confirmed infections versus deaths of COVID-19 in Banglades (as of April 14)

Testing is the only effective window onto the COVID-19 pandemic and how it is spreading. When disease becomes pandemic, testing for it early leads to quick identification of cases, quick treatment for those people and immediate isolation to prevent spread and to trace their contacts. Early testing also helps to identify anyone who came into contact with infected people so they too can be quickly treated. China, South Korea, Taiwan have followed this procedure to use it as one of our most important tools in the fight to slow and reduce the spread and impact of the virus. As expected, very strong positive correlation is found between the number of tests conducted in last 24 hours and the reported infections in Bangladesh (see Figure 4 (a)). The correlation coefficient is 0.96 based on the data reported as on April 14, 2020. As on April 14, Bangladesh has tested only 13,128 case samples which is in a rate 80 per milion and which is much lower than many countries (Max Roser & Ortiz-Ospina, 2020).

**Figure 4:**
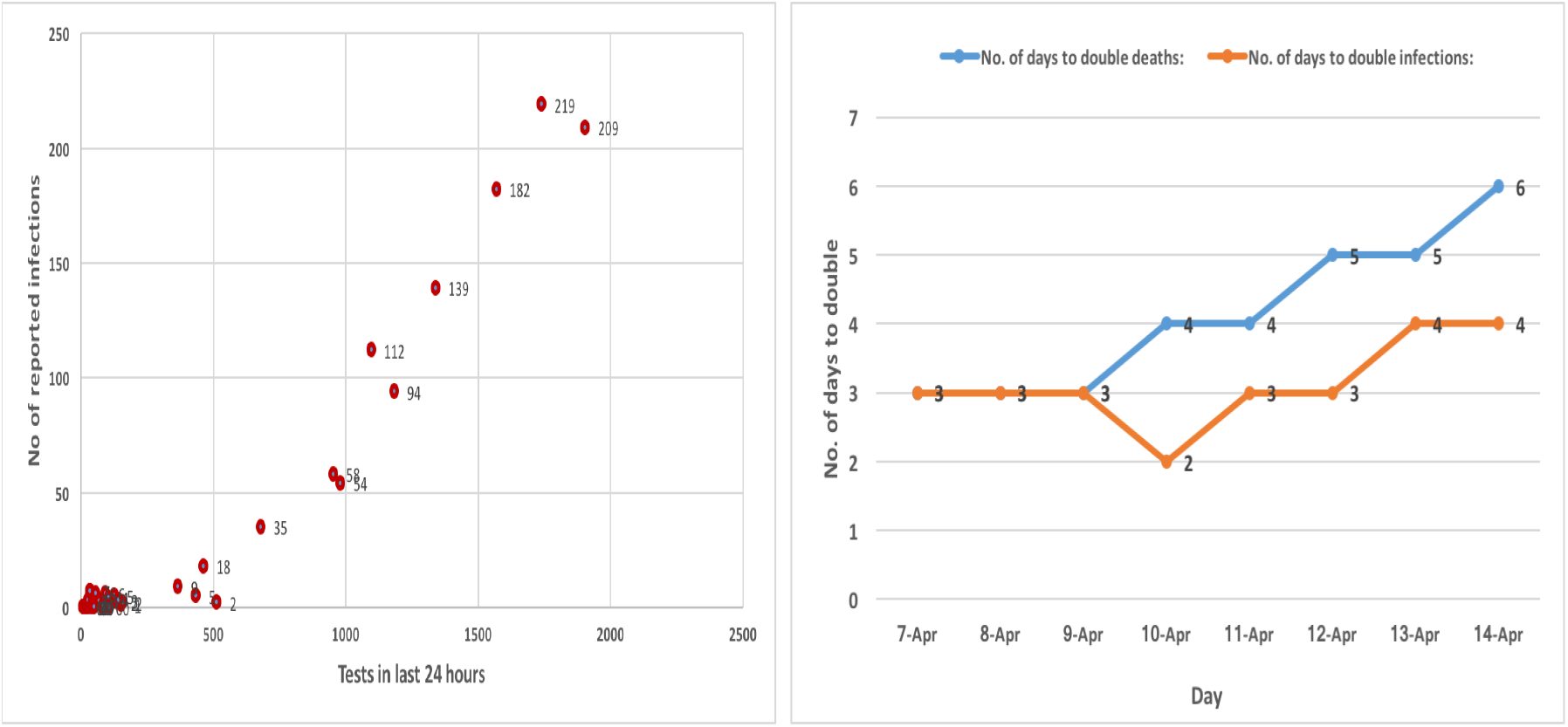
(a) Scatter plot between number of tests and repoted infections of COVID-19 in Banglades (as of April 14) (b) Number of days to double the infections and deaths between April 7 and 14, 2020.

### 4.2 Projection based on Infection Trajectory-Pathway Strategy

Bangladesh is one of the countries who have passed the threshold of 1000 confirmed cases, with many more countries on the cusp (Max Roser & Ortiz-Ospina, 2020). We have presented in Figure 5, the infection trajectory for the countries, based on the data reported as on April 14, who have crossed the 100 case mark and have experience, being well ahead Bangladesh, the pathways that Bangladesh is likely to experience in future. By comparing such trajectories, we would be able to see a clearer picture of how quickly the virus may spread in future in Bangladesh who is on the same track but well behind the track and also how quickly the virus spread within the countries who are on the same trajectory line. We call this procedure as the ITPS (Infection Trajectory-Pathway Strategy). Bangladesh crossed 100 case mark on April 7 exactly one month after the first case that was identified on April 8. However, Figure 5 shows that Bangladesh could follow the same pathway of infection trajectory (between 3 and 5 days to double the cases) that is experienced by any of the five countries-Austria who is 29 days ahead, Isreal who is 25 days ahead, Netherlands who is 31 days ahead, France who is 38 days ahead and United Kingdom who is 32 days ahead. These lead to the projected infections for Bangladesh as reported in Table 1. This table also shows the projected deaths estimated based on the global CFR value which is about 6 (Johns Hopkins University, 2020b) as on April 14, 2020. This projection method suggests that Bangladesh could cross 14,000 case mark and 850 deaths, in worst scenario, by May 13 while infection and death toll could cross 100,000 and 8500 respectively around in May 20.

**Table 1:**
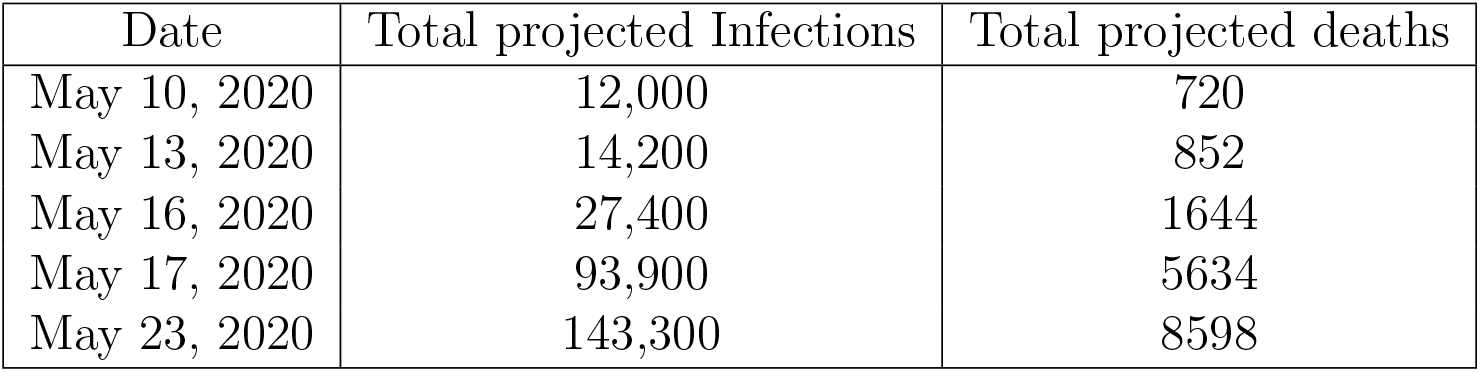
Projected infections and deaths based on ITPS method

| Date | Total projected Infections | Total projected deaths |
| --- | --- | --- |
| May 10, 2020 | 12,000 | 720 |
| May 13, 2020 | 14,200 | 852 |
| May 16, 2020 | 27,400 | 1644 |
| May 17, 2020 | 93,900 | 5634 |
| May 23, 2020 | 143,300 | 8598 |

**Figure 5:**
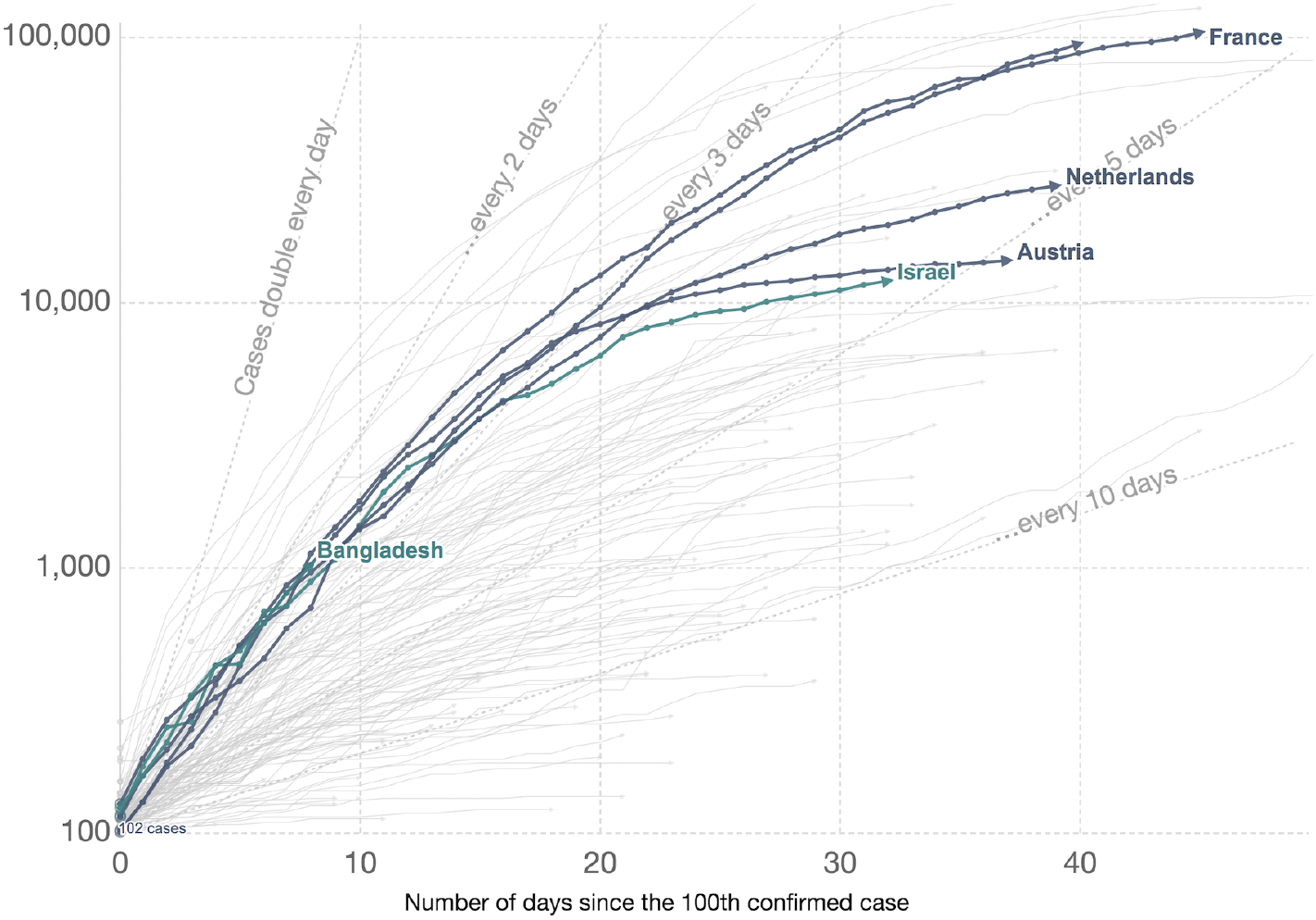
Infection trajectory Bangladesh, Austria, Isreal, Netherlands, France and United Kingdom since 100 confirmed Cases.

### 4.3 Projection based on Polynomial Regression Model

The projections for infected people, severe patients, ICU patients and deaths for short term have been made using the polynomial regression. Table 2 shows the projected numbers with their 95% confidence intervals. According to (Phua et al., 2020), 12% of all reported cases need ICU admissions while 13.4% of all patients are treated as severe. All patientss who are severe need hospital admissions, in addition to some other symptomatice patients with shown sufficient symptoms. In this paper, we have predicted the total infections for short period of time i.e. from April 15 to April 30, 2020 since the model is reasonably good for capturing short term trends. However, the prediction for severe and ICU patients has been carried out based on the findings in (Phua et al., 2020). The deaths are projected from the infected people by assuming that Bangladesh will follow the similar global case fatality rate which is stated as almost 6 as of April 14, 2020 (Max Roser & Ortiz-Ospina, 2020).

**Table 2:** Projected infected people, severe patients, ICU patients and deaths of COVID-19 in Bangladesh by polynomial regression method

| Date | Projected total infections | 95% CI for total infections |  | Projected total severe patients | 95% CI for total severe patients |  | Projected total ICU patients | 95% CI for total ICU patients |  | Projected total deaths | 95% CI for total deaths |  |
| --- | --- | --- | --- | --- | --- | --- | --- | --- | --- | --- | --- | --- |
|  |  | Lower | Upper |  | Lower | Upper |  | Lower | Upper |  | Lower | Upper |
| 15-Apr | 1152 | 1075 | 1230 | 154 | 144 | 165 | 138 | 129 | 148 | 69 | 65 | 74 |
| 16-Apr | 1349 | 1260 | 1438 | 181 | 169 | 193 | 162 | 151 | 173 | 81 | 76 | 86 |
| 17-Apr | 1562 | 1458 | 1665 | 209 | 195 | 223 | 187 | 175 | 200 | 94 | 87 | 100 |
| 18-Apr | 1790 | 1669 | 1912 | 240 | 224 | 256 | 215 | 200 | 229 | 107 | 100 | 115 |
| 19-Apr | 2035 | 1893 | 2177 | 273 | 254 | 292 | 244 | 227 | 261 | 122 | 114 | 131 |
| 20-Apr | 2296 | 2131 | 2460 | 308 | 286 | 330 | 276 | 256 | 295 | 138 | 128 | 148 |
| 21-Apr | 2572 | 2382 | 2763 | 345 | 319 | 370 | 309 | 286 | 332 | 154 | 143 | 166 |
| 22-Apr | 2865 | 2646 | 3083 | 384 | 355 | 413 | 344 | 318 | 370 | 172 | 159 | 185 |
| 23-Apr | 3173 | 2925 | 3422 | 425 | 392 | 459 | 381 | 351 | 411 | 190 | 176 | 205 |
| 24-Apr | 3498 | 3216 | 3779 | 469 | 431 | 506 | 420 | 386 | 453 | 210 | 193 | 227 |
| 25-Apr | 3838 | 3522 | 4155 | 514 | 472 | 557 | 461 | 423 | 499 | 230 | 211 | 249 |
| 26-Apr | 4195 | 3841 | 4548 | 562 | 515 | 609 | 503 | 461 | 546 | 252 | 230 | 273 |
| 27-Apr | 4567 | 4175 | 4960 | 612 | 559 | 665 | 548 | 501 | 595 | 274 | 251 | 298 |
| 28-Apr | 4956 | 4522 | 5390 | 664 | 606 | 722 | 595 | 543 | 647 | 297 | 271 | 323 |
| 29-Apr | 5360 | 4882 | 5838 | 718 | 654 | 782 | 643 | 586 | 701 | 322 | 293 | 350 |
| 30-Apr | 5780 | 5257 | 6304 | 775 | 704 | 845 | 694 | 631 | 756 | 347 | 315 | 378 |

Our projection believes that total infected people and deaths in the Bangladesh will be more than 5700 and almost 350 respectively by the end of April, while the number of severe and ICU patients will be 775 and 695 respectively. Those figures are estimated to be 2296, 138, 308 and 276 respectively by April 20. We have not considered the relationship between deaths and country’s intensive care bed capacity in calculating the potential deaths in our projection, But we believe that the actual projected deaths could be higher than the predicted numbers since the Bangladesh will not have enough beds and intensive care unit (ICU) beds to meet demand [“The Daily DhakaTribune” (March 21, 2020), Nafseen (2018)].

## 5 Discussions and Conclusions

This paper presented the basic analysis results of the current COVID-19 situations in Bangladesh. The paper also proposed an ad-hoc projection strategy known as ITPS (Infection Trajectory-Pathway Strategy) for projecting total infections and deaths suiitable for a moderated period of time like one month or one and half months. The ITPS is a new of its kind in predicting infectee people while it assumes the projected country will follow the similar infection trajectory line or indirectly the similar growth rates of the countries who are already in advanced level of common experiences like infectios and deaths. As reported in Table 1, according to ITPS Bangladesh will have similar number of infections like 14,200 by Austria who is 29 days ahead, 12,000 by Isreal who is 25 days ahead, 27,400 by Netherlands who is 31 days ahead, 143,300 by France who is 38 days ahead and 93,900 by United Kingdom who is 32 days ahead. This paper also presented the projection results by the polynomial regression model which is suitable for short period of time. As reported in Table 2, Bangladesh will cross 3000 infection cases and 190 deaths mark by April 23, while the country will cross 5000 cases and almost 300 deaths mark by April 28. The number of severe and ICU patients will exceed the 560 and 500 marks by April 26, which by the end of April will be 775 and 694 respectively.

The world grapples with the containment of the COVID-19 outbreak but Bangladesh may not be doing so as the number of tests done as of April 14 is over just 13,000 which is not a good number compared to even neibourhood countries such as India, Pakistan. Despite having very strong positive correlation (0.96) between the daily tests and number of infected people, the growth rate of tests in Bangladesh is considerably low. Capacity of tests needs to increase rapidly for detecting the infected people within quick time. This will help to restrict the spread by isolating the infected people and quaranting the susceptible people.

Our projected number of severe and ICU patients might help the government of Bangladesh to prepare the number of hospital beds including ICU beds, health staff to tackle the potential demand of COVID-19 patients. However, our findings provide an indication of the challenges that the Bangladesh health care system will face if the COVID-19 epidemic progresses unabated. For giving tratment to critical patients fully functional ICU beds are crucial. The ICU beds are not useful in the absence of an adequate number of trained critical health-care workers, medical supplies like personal protective equipment (PPE) that are needed for crisis management. Therefor, the government, hospital administrators, and policy makers must work with ICU doctors and nurses to prepare for a substantial increase in critical care bed capacity. Goverment must protect in unprecedented ways the health-care workers from nosocomial transmission and physical exhaustion including several mental health issues. We believe this study will help the Government and health-care workers in preparing their plans for the next two or three weeks. Our predictions by both methods are suitable for both short-term and medium-term interval, and these models can be tuned for forecasting in long-term intervals.

## Data Availability

The data can be obtained from the Novel Coronavirus COVID-19 (2019-nCoV) Data Repository by Johns Hopkins CSSE.

https://github.com/CSSEGISandData/COVID-19

## Author’s Contributions

MHRK carried out the statistical analysis and contributed to draft the manuscript. AH arranged the datasets and contributed to finalize the manuscript.

## References

Boldog, P., Tekeli, T., Vizi, Z., Dnes, A., Bartha, F. A., & Rst, G. (2020). Risk assessment of novel coronavirus COVID-19 outbreaks outside China. Journal of Clinical Medicine, 9 (2), 571.

Chinazzi, M., Davis, J. T., Ajelli, M., Gioannini, C., Litvinova, M., Merler, S., & Vi-boud, C. (2020). The effect of travel restrictions on the spread of the 2019 novel corona-virus (COVID-19) outbreak. Science.

The Daily DhakaTribune. (March 21, 2020).

Dong, D. H. E., & Gardner, L. (2020). An interactive web-based dashboard to track COVID-19 in real time. The Lancet Infectious Diseases.

Grasselli, G., Pesenti, A., & Cecconi, M. (2020). Critical care utilization for the COVID-19 outbreak in Lombardy, Italy: early experience and forecast during an emergency re-sponse. JAMA.

Howard, L. J. (1943). Fitting polynomial trends to seasonal data by the method of least squares. Journal of the American Statistical Association, 38 (224), 453–465.

Hui, D. S., Azhar, E., & Madani, T. A. e. (2020). The continuing 2019-nCoV epidemic threat of novel coronaviruses to global health-the latest 2019 novel coronavirus outbreak in wuhan, china. International Journal of Infectious Diseases, 91, 264–266.

IECDR. (2020). Institute of Epidemiology Disease Control And Research. https://www.iedcr.gov.bd., Accessed April 13.

IHME COVID-19 health service utilization forecasting team. (2020). Forecasting COVID-19 impact on hospital bed-days, icu-days, ventilator-days and deaths by us state in the next 4 months. MedRxiv, DOI: 10.1101/2020.03.27.20043752.

Islam, M. M., Islam, M. M., Hossain, M. J., & Ahmed, F. (2020). Modeling risk of infectious diseases: a case of coronavirus outbreak in four countries,. MedRxiv, DOI: 10.1101/2020.04.01.20049973.

Johannes, L. (2008). Smoothing time series with local polynomial regression on time. Communications in Statistics - Theory and Methods, 37 (6), 959–971.

Johns Hopkins University. (2020a). ”Coronavirus Map”. https://coronavirus.jhu.edu/map.html..

Johns Hopkins University. (2020b). Novel Coronavirus COVID-19 (2019-nCoV) Data Repository by Johns Hopkins CSSE. https://github.com/CSSEGISandData/COVID-19.

Kennedy, P., & Pronovost, P. (2006). Shepherding change: how the market, healthcare providers, and public policy can deliver quality care for the 21st century. Crit Care Med., 34 (3 Suppl), S1.

Kucharski, A. J., Russell, T. W., Diamond, C., Liu, Y., Edmunds, J., Funk, S., & Da-vies, N. (2020). Early dynamics of transmission and control of COVID-19: a mathemati-cal modelling study. The Lancet Infectious Diseases.

Lourenco, J., Paton, R., Ghafari, M., Kraemer, M., Thompson, C., Simmonds, P., … Gupta, S. (2020). Fundamental principles of epidemic spread highlight the immediate need for large-scale serological surveys to assess the stage of the SARS-CoV-2 epidemic. MedRxiv.

Max Roser, H. R., & Ortiz-Ospina, E. (2020). Coronavirus Disease (COVID-19) ? Statistics and Research. Our World in Data. (https://ourworldindata.org/coronavirus)

Ministry of Health and Family Welfare. (2017). Annual Report HSD-2016-17. Government of the People’s Republic of Bangladesh.

Nafseen, M. (2018). Critical Care Medicine: Bangladesh Perspective. Adv J Emerg Med., 2 (3), e27.

Pandeya, G., Chaudharya, P., Guptab, R., & Palc, S. (2020). Seir and regression model based COVID-19 outbreak predictions in india. Arxiv.

Paul, A., Chatterjee, S., & Bairag, N. (2020). Prediction on Covid-19 epidemic for different countries: Focusing on South Asia under various precautionary measures. MedRxiv, DOI: 10.1101/2020.04.08.20055095.

Phua, J., Weng, L., Ling, L., Egi, M., Lim, C.-M., Divatia, J. V., … Du, B. (2020). Intensive care management of coronavirus disease 2019 (COVID-19): challenges and recommendations. Lancet Respir Med..

Roosa, K., Lee, Y., Luo, R., Kirpich, A., Rothenberg, R., Hyman, J. M., & Chowell, G. (2020). Real-time forecasts of the COVID-19 epidemic in China from February 5th to February 24th. Infectious Disease Modelling, 5, 256–263.

WB. (2020). The World Bank data. https://data.worldbank.org/indicator/sh.med.beds.zs, Accessed April 13.

WHO. (2020). Coronavirus disease (COVID-2019) situation reports. Situation report, 84, Accessed April 13.

Wikipedia. (2020). https://en.wikipedia.org/wiki/coronavirus., Accessed April 7.

Xie, J., Tong, Z., Guan, X., Du, B., Qiu, H., & Slutsky, A. S. (2020). Critical care crisis and some recommendations during the COVID-19 epidemic in china. Intensive Care Med, DOI: 10.1007/s00134-020-05979-7.

